# Common Genetic Variation Important in Early Subcortical Brain Development

**DOI:** 10.1101/2022.08.11.22278677

**Authors:** Harriet Cullen, Konstantina Dimitrakopoulou, Hamel Patel, Charles Curtis, Dafnis Batalle, Oliver Gale-Grant, Lucilio Cordero-Grande, Anthony Price, Joseph Hajnal, A David Edwards

## Abstract

1.

Recent genome-wide association studies have identified numerous single nucleotide polymorphisms (SNPs) associated with subcortical brain volumes. These studies have been undertaken in largely adult cohorts. To better understand the role of genetic variability in foetal and perinatal brain development, we investigate how common genetic variation affects subcortical brain development in a cohort of 208 term-born infants from the Developing Human Connectome Project.

We examine six SNPs, that have shown robust associations with subcortical brain volumes in adult studies and ask if these associations exist at birth. We then investigate whether genome-wide polygenic scores (GPSs) for adult subcortical brain volumes are predictive of the corresponding neonatal brain volume. Finally, we explore the relationship between GPSs for psychiatric disorders and subcortical brain volume at birth.

We find the association between SNP rs945270 and putamen volume, seen in adults, is present at birth (p=3.67×10^-3^, β=0.13, SE=0.04). The associations between SNP rs61921502 and hippocampal volume and SNP rs11111090 and brainstem volume are also nominally present in our neonatal cohort. We show that neonatal hippocampal, brainstem, putamen and thalamic volume are all significantly associated with the GPSs for their corresponding volume in adults. Finally, we find that GPSs for five psychiatric disorders and a cross-disorder score are not significantly predictive of subcortical brain volumes or total brain volume at birth. Our results indicate that SNPs important in shaping adult subcortical brain volume are also significant in foetal and perinatal brain development.

**Key Points:**

- We show that the association between the single nucleotide polymorphism, rs945270 and putamen volume, seen in adults, is present in neonates.
- We show that neonatal hippocampal, putamen, brainstem and thalamic volumes are all significantly predicted by the genome-wide polygenic scores for corresponding adult brain volumes.
- We do not find any robust association between genome-wide polygenic scores for psychiatric disorders and neonatal brain volume although we observe several nominal associations.

## 2. Introduction

Imaging-genetic studies in largely adult cohorts have identified many genome-wide significant associations between MRI brain-imaging phenotypes and single nucleotide polymorphisms(1)(2)(3). However, the extent to which common genetic variation, known to be important in shaping adult brain volume, varies over the life course, is still not well understood. Some studies suggest minimal overlap between genetic variants impacting brain volume at different ages (4), whilst others imply more concordance (2)(5). Understanding the role of common genetic variation in shaping early brain development is important; foetal and early neonatal brain development represents a critical phase, dependant on the precise regulation of gene expression(6).

Several recent adult studies have focussed on subcortical brain volumes (2)(3)(7) identifying single nucleotide polymorphisms (SNPs) important in influencing these structures. Differences in the subcortical brain and its related circuitry are thought to be associated with cognitive function and psychiatric and movement disorders (8)(9)(10)(11). The subcortical brain is also particularly vulnerable to abnormal development in infants born preterm (12)(13), abnormalities which are associated with adverse neurocognitive outcome (14)(15).

This study builds on existing work in adult and paediatric populations to explore genetic variation important during foetal and perinatal brain development. We focus on the subcortical brain where robust associations have already been established in adults. Using data form the Developing Human Connectome Project (www.developingconnectome.org)(16) which provides high quality neonatal brain imaging and genome-wide SNP data, we ask if key subcortical SNP-volume associations, present in adults, exist at birth. We then look at whether genome-wide polygenic scores (GPSs), computed from adult genome-wide association studies (GWAS) of the subcortical brain are predictive of the corresponding neonatal brain volume. Finally, we explore the relationship between GPSs for five psychiatric disorders and a cross-disorder score and subcortical and total brain volume at birth.

## 3. Methods

### 3.1 Sample Characteristics

Data for this study were obtained as part of the Developing Human Connectome Project (dHCP) (http://www.developingconnectome.org/). This project has received UK NHS research ethics committee approval (14/LO/1169, IRAS 138070), and written informed consent was obtained from parents.

This study includes infants with genetic data genotyped in the first phase of genetic analysis for the dHCP project and their accompanying imaging data. Individuals with marked cerebral pathology in the subcortical brain were excluded (1 infant). The full mixed-ancestry cohort comprises 418 infants born after 37 weeks of gestation with accompanying MRI and genetic data and includes infants of all ancestries. A subsample of European-ancestry individuals, which formed the principal cohort for our investigation, comprised 208 infants. Infants were typically scanned within the first few weeks of life (range: zero weeks to five weeks and six days). Our study therefore captures the effects of brain development predominantly during the foetal period but also includes a brief perinatal period, dependant on the precise timing of the infant’s scan. Sample characteristics for the full mixed-ancestry cohort the European-Asian ancestry cohort and the European ancestry cohort are detailed in Table 1.

**Table 1.** Summary Statistics for three neonatal cohorts. Values indicated are mean and standard deviation.

|  | European-Ancestry Cohort | European - Asian Ancestry Cohort | Full Mixed-Ancestry Cohort |
| --- | --- | --- | --- |
| Number of Infants (Male, Female) | 208 (110, 98) | 258 (139,119) | 418 (228,190) |
| Mean Gestational Age at Birth (weeks) | 40.1 ± 1.2 | 40.1 ± 1.3 | 40.0 ± 1.2 |
| Mean Postmenstrual Age at Scan (weeks) | 41.5 ± 1.7 | 41.5 ± 1.7 | 41.2 ± 1.7 |
| Mean Intracranial Volume (cm<sup>3</sup>) | 456 ± 62 | 452 ± 61 | 444 ± 60 |

### 3.2 Imaging Data

T1 and T2 images were acquired at term-equivalent age on a 3-Tesla Philips scanner with a dedicated neonatal imaging system (17). Volume estimates for amygdala, hippocampus, brainstem, caudate and thalamus bilaterally and total brain volume were generated using the dHCP neonatal structural pipeline (18). Volume estimates for the bilateral volume of the putamen and pallidum, which are not available from the dHCP structural pipeline, were extracted using a neonatal version (19) of the Automated Anatomical Labelling (AAL) atlas adapted to a high-resolution dHCP template (20). AAL parcellation propagation was performed as previously described in (21).

### 3.3 Genetics Data

Infant saliva was collected using the Oragene DNA OG-250 kit, DNA was extracted and genotyped for SNPs genome-wide on the Illumina Infinium Omni5-4 v1.2 array. Basic quality control was performed according to the pipeline (22). Further quality control was then undertaken, this included the removal of duplicate samples and the removal of individuals with discordant gender information or individuals with a genotyping failure of more than 1% of SNPs.

SNPs were excluded that had a minor allele frequency <= 0.05, were missing in > 1% of individuals or deviated from Hardy-Weinberg equilibrium with a *P* value < 1 × 10^-5^. Non-autosomal SNPs were removed. Further, individuals were excluded if there was genetic evidence of relatedness (pi_hat >= 0.1875). In such cases one member of each pair was randomly retained.

The dataset was imputed to the Haplotype Refence Consortium reference panel (23) on the Michigan Imputation Server. The VCF files returned were converted to plink files using a genotype calling threshold of 0.9. The imputed markers underwent a second stage of quality control. SNPs were excluded that had a minor allele frequency < = 0.05, were missing in more than 1% of individuals, deviated from Hardy-Weinberg equilibrium with a *P* value < 1 × 10^-5^ or had an imputation Rsq value of less than or equal to 0.8. All quality control was performed with PLINK 1.9 (https://www.cog-genomics.org/plink2).

Ancestry sub-populations were identified by merging our cohort with 2504 individuals from the 1000 Genomes Project using a subset of common autosomal SNPs (24). We then performed a principle-component (PC) analysis in plink and examined sequential PC plots utilizing population labels provided for individuals in the 1000 Genomes Project to manually identify subpopulations within our sample.

### 3.4 Single SNP-volume Analysis

#### 3.4.1 Selection of SNP-volume Pairs

The starting point for this work were the results of Satizabal et al. (2019) (3) who explored SNP-volume associations for the accumbens, amygdala, brainstem, caudate nucleus, globus pallidus, putamen and thalamus using GWAS. Their cohort included almost 40,000 individuals from the Cohorts of Heart and Aging Research in Genomic Epidemiology (CHARGE) consortium, the Enhancing Neuro Imaging Genetics through Meta-Analysis (ENIGMA) consortium and UK Biobank. The hippocampus was not included as it had been recently studied using 33536 individuals from the CHARGE and ENIGMA consortiums (7).

To identify a set of SNP-volume pairs to explore in our neonatal sample, we first selected the most robust association for each subcortical volume from either (3) or (7). We then looked at whether these results had been replicated, for a comparable phenotype (*P* < 5×10^-8^), in the recent UK Biobank study (1) which performed GWASs for multiple brain-imaging phenotypes, including subcortical brain volumes, in 8428 individuals. For the hippocampus, we included the two most significant associations because of the low minor allele frequency of the most robust association. Details of the SNP-volume pairs we considered, their associated *P* values in the original studies (3)(7), and whether they were replicated in (1) are given in Supplementary Table 1. The nucleus accumbens was not included because this volume was not extracted using our neonatal atlases. Our final list included six SNP-volumes pairs.

#### 3.4.2 Statistical Analysis of SNP-Volume Pairs

We investigated the SNP-volume associations in our neonatal cohort using the same approach as the adult studies in which the associations were identified (1)(3)(7). The additive SNP dosage value was regressed against the bilateral subcortical brain volume of interest. We included gestational age at birth, postmenstrual age at scan, sex, intracranial volume and ancestry principal components as covariates. The number of ancestry principal components included was dependant on the cohort under investigation: three, five and nine were used respectively for the European ancestry, European-Asian ancestry and full mixed-ancestry cohorts respectively.

The main analysis was undertaken in our European-ancestry cohort (n=208). Six SNP-volume pairs were explored with a Bonferroni-corrected *P* value for association of *P* < 0.05/6 = 0.0083. To make the best use of the ancestral diversity in our dHCP dataset, SNP-volume associations reaching nominal significance in the European-ancestry cohort were also explored in the larger European - Asian ancestry and the full mixed-ancestry cohorts.

### 3.5 GPSs for Adult Subcortical Brain Volumes and Neonatal Brain Volume

The first part of this study explored single genome-wide significant SNP-volume associations in our neonatal cohort. We then asked whether broader genetic variation, associated with adult subcortical brain volumes, predicts morphological differences in the infant brain. We computed GPSs for seven subcortical brain volumes using summary statistics available from the work of (3) and (7) in our European-ancestry cohort and explored the association of these scores with the corresponding neonatal brain volume.

GPSs are the sum of an individual’s risk alleles, weighted by the allele’s effect size, which is estimated from the GWAS for the relevant trait or phenotype. GPSs were computed in PRSice-2 (25) which uses a subset of SNPs, extracted following *P-*value informed clumping, that exceed a specified GWAS *P-*value threshold. Scores were computed for our neonatal cohort at six *P-*value thresholds (1×10^-8^, 1×10^-6^,0.001, 0.01, 0.1,1). The 1000 Genomes project was used as an external linkage disequilibrium reference dataset (24).

We used a linear regression model to explore possible association between infant subcortical brain volume and GPS for adult subcortical brain volume, including sex, gestational age at birth, postmenstrual age at scan, intracranial brain volume and three ancestry principal components as covariates. Our principal aim was to assess whether the GPS for a given adult subcortical brain volume predicts the corresponding neonatal brain volume. Seven subcortical GPSs, computed at six *P-*value thresholds were compared with the corresponding neonatal brain volume. Both the neonatal brain volumes and the differently thresholded GPSs are highly correlated. We used the method proposed by (26) to compute the effective number of independent tests performed, accounting for the correlation structure between measures, giving a corrected *P* value of *P* < 4.16×10^-3^. Additional results looking at nonspecific associations between all adult subcortical GPSs and all neonatal brain volumes are presented in Supplementary Figure 1.

### 3.6 GPSs for Psychiatric Disorders and Neonatal Brain Volume

Finally, we investigated whether common genetic variation associated with psychiatric disorders is predictive of early morphological differences in the subcortical brain or total brain volume. We computed GPSs for autism spectrum disorder (ASD), attention deficit hyperactivity disorder (ADHD), schizophrenia, bipolar disorder, major depressive disorder and cross-disorder (27) (including eight psychiatric disorders: anorexia nervosa, ADHD, ASD, bipolar disorder, major depression, obsessive-compulsive disorder, schizophrenia, and Tourette syndrome) in our European-ancestry cohort and explored the relationship with seven subcortical brain volumes and total brain volume. Details of the summary statistics used to compute psychiatric GPSs are given in Supplementary Table S2.

GPSs were generated in PRSice-2 (25) as previously described. We used a linear regression model to explore a possible association between neonatal brain volume and psychiatric GPS, including sex, gestational age at birth, postmenstrual age at scan, intracranial brain volume and three ancestry principal components as covariates. For the analysis with total brain volume, sex, gestational age at birth, postmenstrual age at scan and three ancestry principal components were included as covariates. Six psychiatric GPSs were considered at six *P-* value thresholds for seven subcortical volumes and total brain volume. Using the method proposed by (26) to compute the effective number of independent tests performed gave a corrected *P* value for association of *P* < 9.26×10^-4^ across all six disorders or *P* < 5.56×10^-3^ within a single psychiatric disorder.

## 4. Results

### 4.1 Single SNP-Volume Analysis

Results of the SNP-volume association analysis are detailed in Table 2. There was a statistically significant association between the number of C alleles at SNP rs945270 and putamen volume in our neonatal cohort: a greater number of C alleles was associated with a larger volume (β =0.128, SE=0.044, *P*=3.67×10^-3^). There was a nominal association (*P* < 0.05) of SNP rs61921502 with hippocampal volume (β =-0.106, SE=0.047, *P*=0.026) and SNP rs11111090 with brainstem volume (β =-0.072, SE=0.036, *P*=0.042). For all significant and nominally significant results the direction of association was the same as in previous adult studies (1)(3)(7). There was no evidence in our neonatal cohort for the SNP-volume associations for the pallidum and caudate, previously identified in adults.

**Table 2.** SNP-volume results for the European-ancestry (n=208) cohort Results of association analyses of the six selected SNP-volume pairs in our cohort of 208 European-ancestry neonates. Allele 1 is the coded allele; Allele 2 is the non-coded allele. Standardized beta coefficients (β) and standard errors (SE) are given with respect to Allele 1. The table provides raw *P* values. Results surviving Bonferroni-correction (*P* <0.0083) are indicated in bold. Results indicating nominal significance (*P* <0.05) are indicated in italics.

| Volume | Marker | Allele 1 | Allele 2 | Allele 2 AF | Allele 1 AF | Rs<sub>q</sub> | European ancestry cohort (n=208) |  |  |
| --- | --- | --- | --- | --- | --- | --- | --- | --- | --- |
|  |  |  |  |  |  |  | P value | β | SE |
| **Putamen** | **rs945270** | **C** | **G** | **0.51713** | **0.48287** | **0.97845** | **3.67x10<sup>-3</sup>** | **0.1284** | **0.0437** |
| Pallidum | rs2923447 | **G** | T | 0.58867 | 0.41133 | 0.98606 | 0.3725 | 0.0418 | 0.0468 |
| Caudate | rs3133370 | **C** | T | 0.62460 | 0.3754 | Genotyped | 0.6052 | -0.0256 | 0.0495 |
| *Brainstem* | *rs11111090* | **C** | A | 0.62688 | 0.37312 | *0.9914* | *0.0425* | *-0.0723* | *0.0356* |
| Hippocampus | rs77956314 | **C** | T | 0.92469 | 0.07531 | 0.98433 | 0.340 | 0.0459 | 0.0480 |
| *Hippocampus* | *rs61921502* | **G** | *T* | 0.88988 | 0.11012 | *Genotyped* | *0.0262* | *-0.1058* | *0.0472* |

We investigated the SNP-volume associations for the putamen, hippocampus and brainstem in our larger, European-Asian ancestry and full mixed-ancestry cohorts. The putamen result replicated in both the European – Asian ancestry (β = 0.125, SE=0.040, *P* = 1.89×10^-3^) and full mixed-ancestry cohorts (β = 0.122, SE=0.034, *P* = 4.16×10^-4^) suggesting SNP rs945270 may be associated with early putamen development across multiple ancestral groups. The SNP rs61921502 – hippocampal association remained nominally significant in both the European – Asian ancestry (β =-0.101, SE= 0.043, *P* = 0.020) and full mixed-ancestry cohorts (β =-0.071, SE= 0.034, *P* = 0.038), again suggesting the association may extend across different ancestries.

In contrast, whilst the rs11111090-brainstem association remained nominally significant in the European - Asian ancestry cohort (β =-0.071, SE= 0.032, *P* = 0.029), it was not significant in our full mixed-ancestry cohort (*P* = 0.130). The dilution of the SNP – phenotype relationship in ancestrally mixed populations is well documented (28) and may be a result of differential tagging in different populations. For completeness, the additional, non-significant associations were explored in the European - Asian ancestry and full mixed-ancestry cohorts with results detailed in the Supplementary Analysis Tables S3a and S3b respectively.

### 4.2 GPSs for Adult Subcortical Brain Volume and Neonatal Brain Volume

Results are shown in Figure 1. There was a trend of association between all neonatal subcortical brain volumes and the corresponding adult subcortical GPSs; all brain volumes, except for the amygdala, had at least one GPS *P-*value threshold where there was an uncorrected (*P* < 0.05) association with the corresponding adult GPS. Of the seven volumes explored, only the hippocampus, brainstem, putamen and thalamus showed a robust association (*P* < 4.17x 10^-3^) with the adult GPS. These associations were generally seen at a single GPS threshold with other thresholds indicating uncorrected association or no association. Neonatal brainstem volume was most robustly predicted by adult GPS; neonatal brainstem volume was significantly associated with adult brainstem GPS at four of the six *P-* value thresholds explored.

**Figure 1.**
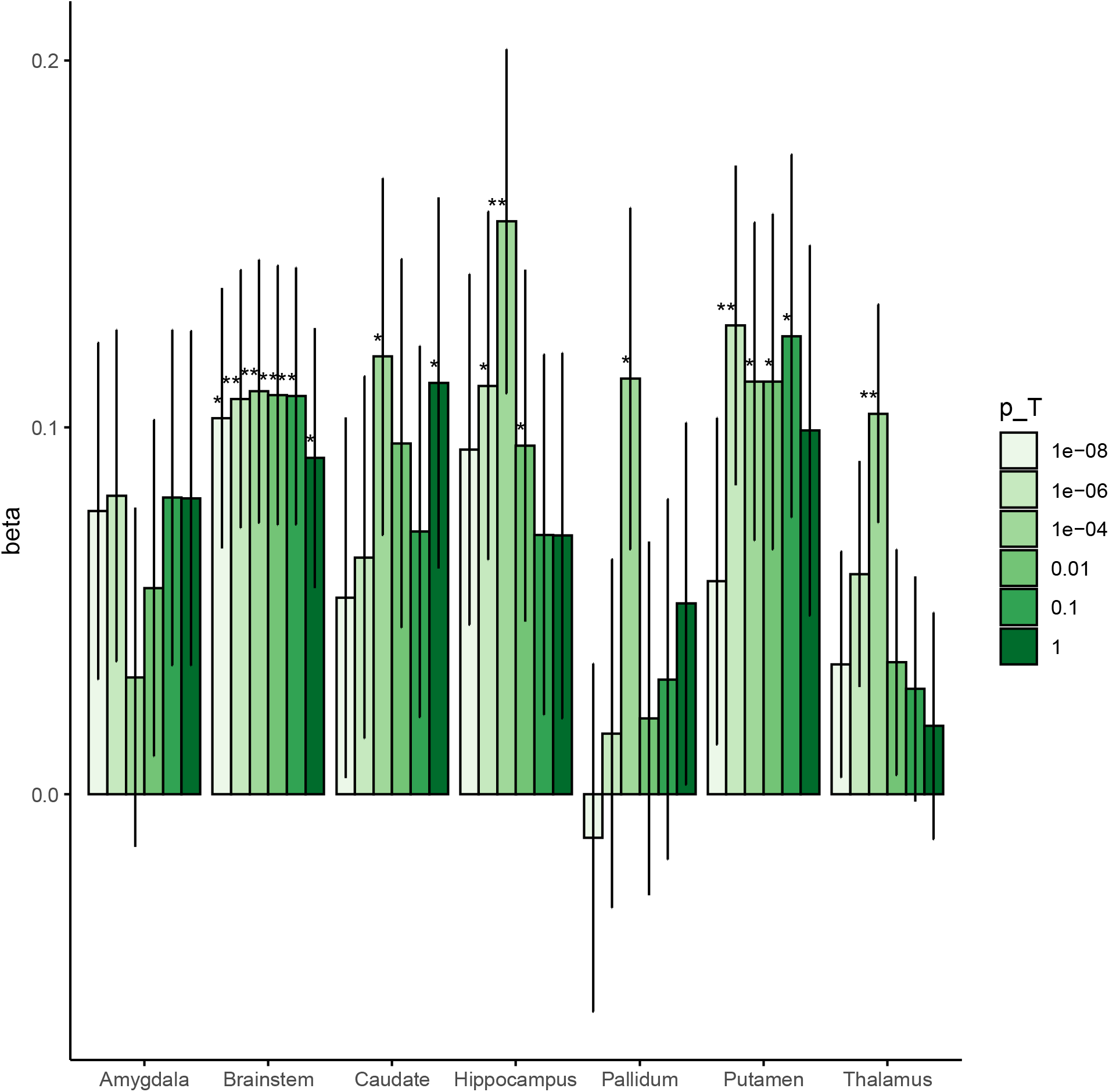
Association between Adult Subcortical GPS and Neonatal Brain Volume The beta coefficients for the association between adult subcortical GPS and the corresponding neonatal brain region. The coloured bars represent the different values for the GWAS *P*-value thresholds investigated for each GPS (p_T_ = 1×10^-8^, 1×10^-6^, 0.0001, 0.01, 0.1 and 1). The error bars indicate standard errors in the beta coefficients. ** Results surviving multiple testing correction (*P* < 4.17×10^-3^), * nominal association (uncorrected P value < 0.05).

For completeness, an investigation of non-region-specific associations was also undertaken the results of which are presented in a series of heatmaps in Supplementary Figure 1. As expected, the most robust associations are the region-specific associations. Adult subcortical GPSs occasionally show uncorrected association with non-region-specific neonatal brain volumes. Of note, the amygdala volume, which is poorly predicted by the amygdala GPS, is significantly predicted by the hippocampal GPS at the most stringent *P-* value threshold (p_T_ = 1×10^-8^). Furthermore, the putamen GPS is also predictive of the neonatal pallidum volume and the caudate GPS of neonatal putamen volume.

### 4.3 GPSs for Psychiatric Disorders and Neonatal Brain Volume

Results are displayed in the heatmap shown in Figure 2. None of the neonatal brain volumes showed an association with psychiatric GPS that survived multiple-testing correction across the six psychiatric GPS considered (*P* < 9.26×10^-4^). Three associations survived the within-disorder testing correction (*P* < 5.56×10^-3^). ADHD GPS was negatively associated with total brain volume (β =-0.125, SE = 0.040, *P*=1.99×10^-3^, p_T_=0.01); greater genetic risk of ADHD was associated with smaller total brain volume. Cross disorder risk was also negatively associated with total brain volume (β = -0.129, SE=0.045, *P*=4.56×10^-3^, p_T_=1). Finally cross-disorder risk also showed a positive association with amygdala volume (β = 0.149, SE=0.046, *P* =1.37×10^-3^, p_T_ =0.01); greater genetic risk for multiple psychiatric disorders was associated with larger amygdala volume.

**Figure 2.**
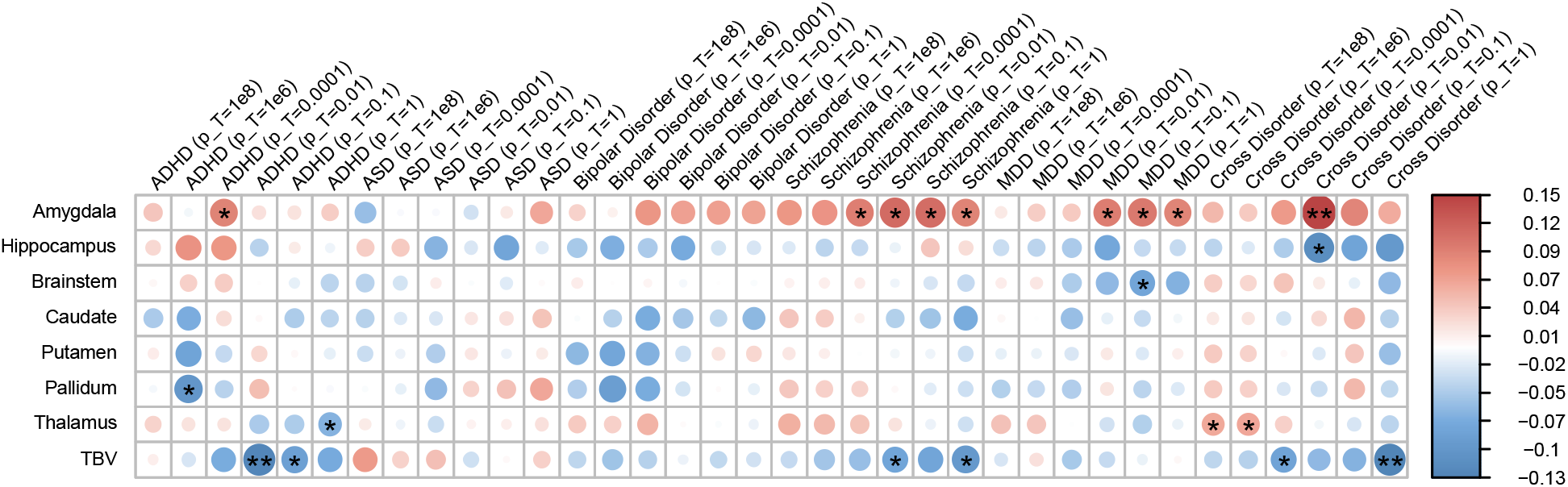
Associations between GPSs for Psychiatric Disorders and Neonatal Subcortical and Total Brain Volume Heatmap for associations between six psychiatric GPSs and seven neonatal subcortical brain volumes and total brain volume (TBV). Associations are shown for six GWAS *P-* value thresholds (p_T_ = 1×10^-8^, 1×10^-6^, 0.0001, 0.01, 0.1 and 1) for each psychiatric GPS. No results survived multiple-testing correction across all psychiatric disorders (*P* < 9.26×10^-4^). ** Results reaching within-disorder significance (*P* < 0.0056). * Results reaching nominal significance (*P* < 0.05). ADHD = Attention Deficit Hyperactivity Disorder, ASD = Autism Spectrum Disorder, MDD = Major Depressive Disorder.

Several additional uncorrected associations for ADHD, schizophrenia, major depressive disorder and cross-disorder with neonatal brain volumes were observed that are detailed as follows. There was an uncorrected positive association of ADHD GPS with amygdala volume (β = 0.093, SE =0.046, *P*=0.045, p_T_=0.0001). In contrast ADHD GPS showed nominal negative associations with both pallidum volume (β = -0.101, SE =0.046, *P*=0.030, p_T_=1×10^-6^) and thalamic volume (β = -0.067, SE= 0.030, *P*=0.027, p_T_=1). The most robust association for ADHD GPS was the negative association with total brain volume reported above.

There was a nominal negative association of schizophrenia GPS with total brain volume (β =-0.096, SE=0.041, *P*=0.020, p_T_=1) and a nominal positive association with amygdala volume (β = 0.113, SE= 0.046, *P*=0.0148, p_T_=0.01). There were nominal associations of major depressive disorder GPS with both amygdala volume and brainstem volume, greater genetic risk for major depressive disorder was associated with larger amygdala volume (β =0.101, SE=0.046, *P*=0.029, p_T_=0.1) but smaller brainstem volume (β = -0.080, SE = 0.036, *P* = 0.027, p_T_ = 0.1).

Finally, cross-disorder GPS indicated a couple of associations that survived within-pathology testing corrections that were detailed above: a positive association with amygdala volume and a negative association with total brain volume. Cross-disorder GPS also indicated a nominal positive association with thalamic volume (β = 0.064, SE= 0.030, *P*=0.037, p_T_=1×10^-6^) and nominal negative association with hippocampus volume (β = -0.113, SE=0.049, *P*=0.021, p_T_=0.011).

Whilst there were no associations between psychiatric GPSs and neonatal brain volume that survived the disorder-wide multiple testing correction, Figure 2 indicates a pattern of positive correlation between most psychiatric GPSs and amygdala volume and a pattern of negative correlation between psychiatric GPSs and total brain volume. In both cases GPS for ASD were an exception to these trends.

## 5. Discussion

This work asked if common genetic variation, known to be important for adult subcortical brain volume, is important in foetal and perinatal brain development. GWAS studies have explored SNP-phenotype associations for multiple brain-imaging phenotypes. Here we focused on subcortical brain volumes which have been studied in large adult cohorts (3)(7) and for which comparable phenotypes could be extracted in our neonatal cohort.

We explored six SNP-volume associations that are well established in adult cohorts. Of these, only the association of rs945270 with putamen volume was significant in our neonatal cohort. This is notably the most significant result identified in adult studies (3)(7). Two further SNP-volume pairs were nominally significant in our cohort: SNP rs61921502 with hippocampal volume and SNP rs11111090 with brainstem volume.

rs945270 is an intergenic locus downstream of the Kinectin 1 (KTN1) gene and has been shown to alter the expression of KTN1 in both blood and brain tissue (29). More specifically Hibar et al. (2) showed that the C allele, which is associated with larger putamen volume, increases expression of KTN1 in the frontal cortex and putamen. Luo et al. (30) showed KTN1 is significantly expressed in the putamen in three adult cohorts and Kang et al. (31) showed that during late foetal development KTN1 is more highly expressed in the thalamus, hippocampus and striatum than in the cortex. KTN1 encodes the protein kinectin, a receptor that allows vesicle binding to kinesin and is involved in organelle transport. Impairment of kinesin has been recognised in neurological diseases with a subcortical component (32) and KTN1 variants have also been associated with ADHD (33)(30).

Our dHCP cohort includes infants of diverse ancestry. This diversity, an asset of this cohort, presents challenges in imaging-genomic studies. The GWAS results we explored, and the summary statistics used to compute the GPSs were from predominantly European-ancestry individuals. For this reason, our analysis was undertaken in a European-ancestry subsample of the dHCP. However, for the single SNP-volume investigations, we were interested to know if the results could be generalisable in a larger, more ancestrally diverse group.

We explored our significant and nominally significant SNP-volume associations in both our European - Asian ancestry and full mixed-ancestry cohorts. Our results showed similar trends to those observed in the larger adult studies. The rs945270-putamen result was consistent across all cohorts and was more robust in our larger European-Asian ancestry and full mixed-ancestry cohorts. Satzibal et al.(3) found the direction of the rs945270 – putamen association was the same for all ethnicities and the strength of association increased in a meta-analysis including different populations.

The results for rs61921502 and rs11111090, both nominally significant in our European-ancestry cohort, were more variable. The SNP rs61921502-hippocampal association remained nominally significant in our European - Asian ancestry and full mixed-ancestry cohorts but was not more robust in these larger groups. In adult studies, Hibar et al (2017) (7) found SNP rs61921502 was borderline associated with hippocampal volume in an African cohort but found no association in the Asian or Mexican-American ancestry cohorts. The SNP rs11111090-brainstem association remained nominally significant in our European - Asian ancestry cohort but was not significant in the full mixed-ancestry cohort. The adult GWAS did not see concordance in the direction of association of SNP rs11111090 with brainstem volume for different ancestries and the meta-analysis including non-European ancestry individuals was less significant than in the European-only ancestry group (3).

Work in the literature has started to explore the role of genetic variation in early brain development. However, the large sample sizes required for hypothesis-free GWAS makes such studies of brain-imaging phenotypes in infants scarce. Xia et al.(4) undertook a GWAS of global brain tissue volumes in 561 infants and compared their results with two large-scale neuroimaging GWAS, one in adolescents and another in adults. They found only minimal overlap between common variants impacting brain volume at different ages. In contrast, in a large, but predominantly adult study, including 30717 individuals aged 9 to 97, Hibar et al.(2) asked if the phenotypic effects of SNPs were related to the age of individuals. Only one of the eight significant loci they identified appeared to have an age-related effect and they argued that most SNP-volume effects, including the rs945270-putamen association, are likely to be stable across the life course. However, their cohort did not include infants or young children.

To complement our work looking at single, genome-wide significant SNPs, we also explored broader genetic variability that might be important in shaping neonatal subcortical brain volume. We asked if GPSs, computed from the summary statistics of adult subcortical volume GWAS, were predictive of the corresponding volumes in our neonatal cohort. A GPS is an estimate of an individual’s genetic liability to a trait or phenotype calculated by summing genome-wide all trait-associated alleles, weighted by their estimated effect size (34). GPSs have been shown to achieve greater predictive power when they include a larger number of SNPs and are not limited to SNPs reaching genome-wide significance (35). As such, GPSs provide a means of probing a broader range of genetic variation that is potentially relevant to early subcortical brain development.

Whilst most adult subcortical GPSs showed a trend of association with the corresponding neonatal brain volume, only the hippocampal, brainstem, putamen and thalamic volumes were robustly predicted by their GPSs. These volumes included the three volumes for which we observed single SNP-volume associations in our cohort. In the supplementary analysis we present results for non-region-specific associations for the subcortical GPSs and neonatal brain volumes. As expected, the non-region-specific associations are few and generally modest (if present).

Our work adds to the emerging literature exploring the role of genetic polymorphisms, known to be important in shaping adult brain morphology, in early brain development. We compare our results with those of Lamballais et al. (5) who computed GPSs for adult subcortical brain volumes and asked how predictive they are of paediatric subcortical brain volume and infant gangliothalamic ovoid diameter (assessed via ultrasound). In their paediatric cohort, imaged at around 10 years, they observed robust associations between all adult GPS scores and the corresponding paediatric subcortical brain volumes. Interestingly, both our study and theirs find the strongest associations for the putamen and brainstem; genetic polymorphisms clearly exist that are important in shaping the morphology of these structures from foetal life through childhood to adulthood.

In general, our subcortical GPS-volume results imply weaker associations than the paediatric sample of (5). This may be a result of our smaller cohort but is also likely influenced by our earlier imaging timepoint. The paediatric brain reaches its maximum size between 10 and 12 years (36) and we know the subcortical brain develops rapidly during early childhood (37). Our neonatal cohort represents a snapshot in the perinatal period soon after birth and foetal development. It is possible that genetic variation plays a less measurable role in shaping the subcortical brain during foetal and perinatal development as compared with later in childhood. It is also possible that the genes and gene pathways most important in influencing subcortical morphology at birth may differ from those in older children and adults.

Studies exploring heritability of brain imaging phenotypes over the life course suggest that heritability may increase slightly from infancy to childhood (38)(39). Furthermore, work looking at genetic variation important in the rate of change of brain structure suggests that heritability is higher in adults than in children implying genetic variation may play a greater role in explaining structural changes as individuals get older. The notion that there may be other important factors relevant early in the life-course is supported by the mediation analysis of Lamballais et al. (5) which suggests that the genetic effects on subcortical volumes during infancy explain only a small part of the volumes in early childhood.

Extensive literature implicates the subcortical brain in the pathology of psychiatric disorders (6). As an extension of our work focused on common genetic variation important in shaping the early subcortical brain, we asked if genetic variation associated with psychiatric disorders is predictive of neonatal subcortical and total brain volume. We found no robust associations between any of the seven subcortical volumes or total brain volume and GPSs for ASD, ADHD, bipolar disorder, schizophrenia, major depressive disorder, or a cross-disorder risk. Like the GWAS studies of brain volumes, much of the work exploring associations between psychiatric GPS and subcortical brain volume has been undertaken in adults. However, in contrast to the brain-volume studies, robust associations have been few.

A large Biobank study(40) looked at the relationship between subcortical brain volume with GPSs for major depressive disorder, bipolar disorder and schizophrenia and found no robust associations, noting only a modest negative association between thalamic volume and schizophrenia risk. Franke et al. (41) used linkage disequilibrium (LD) score regression to explore shared genetic architecture between subcortical brain volume and genetic variability associated with schizophrenia and did not find any overlap. Similarly, no association was found between schizophrenia GPS and subcortical brain volume in a UK biobank sample of 12490 participants (42). Grama et al. (43), also looked at schizophrenia genetic risk and subcortical brain volume and found only one robust association: a negative association with the right pallidum (43). Satizabal et al. (3) used linkage disequilibrium score regression to explore genetic correlation between bipolar disorder, schizophrenia and ADHD and subcortical brain volume; they found an inverse correlation between brainstem volume and genetic liability for ADHD but no associations with bipolar disorder or schizophrenia.

Our study is different from those described above as we explored psychiatric-imaging associations at birth. A few studies have specifically explored psychiatric GPS and brain volume early in the lifecourse. Acosta et al. (44) looked at caudate and putamen volumes in association with risk for major depressive disorder in 105 infants and found greater genetic risk for major depressive disorder associated with smaller caudate volume in female as compred with male infants. Xia et al. (4) looked at genetic liability for psychiatric disorders and brain volume in a cohort aged between 0 and 24 weeks. They found no association between GPS for either schizophrenia or ASD and total grey or white matter volume, intracranial volume, or volume of cerebral spinal fluid. Alemany et al. (45) looked at associations between brain volume and psychiatric GPS in 1139 children imaged at around 10 years of age. They observed no significant associations between GPS for schizophrenia or bipolar disorder but found greater ADHD GPS was associated with smaller caudate and total brain volume. Our data also showed a nominal association of ADHD GPS with total brain volume; like Alemany et al. (45) we found greater ADHD GPS was associated with smaller total brain volume, however, the association with caudate volume was not evident in our cohort. Alemany et al. (45) also found a positive association of ASD PRS with total brain volume which was not seen in our study. It is interesting that we observe the association between ADHD and total brain volume, as seen by Alemany et al. (45) in their paediatric cohort, albeit at a nominal level, and it would be helpful to explore this in future neonatal and paediatric cohorts.

We have previously explored the relationship between subcortical brain volume and psychiatric genetic risk in preterm infants (46). We hypothesised that genetic variability associated with psychiatric disorders might increase vulnerability to abnormal brain development in infants subjected to the environmental stress of prematurity. We showed an association between genetic risk for multiple psychiatric disorders and lentiform nucleus (putamen and globus pallidus) volume: greater genetic risk was associated with smaller volume in preterm infants. We then extended this work, looking at the influence of genetic risk for psychiatric disorders on cognition for both term and preterm infants. Using data from the Twins Early Development Study we showed that individuals with a greater genetic risk burden for either bipolar disorder or schizophrenia appear to be more vulnerable to the adverse effects of prematurity on cognition (47). In our current study, we explore the relationship between psychiatric genetic risk and brain volume in a term-born cohort, distinct from the preterm infants previously studied and find limited evidence for psychiatric GPS – volume associations. If our previous hypothesis, that genetic variation associated with psychiatric disorders plays a more significant role in preterm brain development than in term-born infants is correct, then we might not expect to see significant psychiatric GPS – volume associations in our current cohort. However, the small size of both studies makes it difficult to draw firm conclusions.

A significant limitation of this work is our small sample size. This study was undertaken in a substantially smaller cohort than the adult and paediatric studies with which we compare our results. The primary adult studies used for SNP-volume exploration and from which our GPS for subcortical volumes were computed (3) (7) included upwards of 30,000 individuals. In this work we explored the most significant variants from these studies that were additionally replicated in the UK Biobank study (1) which itself included over 8000 adults and a replication cohort of 930. Despite our limited numbers, we do observe one, previously established SNP-volume association, and show nominal association for two further SNP-volume pairs. Further, we find that GPSs for adult subcortical brain volume are broadly predictive of neonatal subcortical volumes with significant associations for the brainstem, hippocampus, putamen, and thalamus.

## 6. Conclusions

Our results indicate that the common genetic variation important for adult subcortical brain volume plays a significant role in foetal and perinatal brain development. They also suggest that the genetic variability most relevant in shaping neonatal brain morphology, may not be fully represented by the genetic variability shaping adult brain morphology. It is possible that during foetal and perinatal development other factors, including genes and genetic pathways distinct to this period, may play prominent roles. Further analysis with foetal and neonatal cohorts is required to better understand how the genetic mechanisms shaping brain morphology might differ in this early developmental period.

## Data Availability

The dHCP is an open‐access project. The imaging and demographic data used in this study can be downloaded by registering at https://data.developingconnectome.org/

## Acknowledgements

The authors acknowledge use of the research computing facility at King’s College London, Rosalind (https://rosalind.kcl.ac.uk).

## Notes

**Funding statement:** The Developing Human Connectome Project was funded by the European Research Council under the European Union Seventh Framework Programme (FP/20072013)/ERC Grant Agreement no. 319456. This work was supported by the NIHR Biomedical Research Centres at Guys and St Thomas NHS Trust and the South London and Maudsley NHS Trust; the ESPRC/Wellcome Centre for Medical Engineering at Kings College London ((WT 203148/Z/16/Z); and the MRC Centre for Neurodevelopmental Disorders. The views expressed are those of the authors and not necessarily those of the NHS, the National Institute for Health Research or the Department of Health. The funders had no role in the design and conduct of the study; collection, management, analysis, and interpretation of the data; preparation, review, or approval of the manuscript; and decision to submit the manuscript for publication.

DB received support from a Wellcome Trust Seed Award in Science [217316/Z/19/Z]. LCG received support from the Comunidad de Madrid-Spain Support for R&D Projects [BGP18/00178]. H.C is an academic clinical lecturer in Clinical Genetics at Kings College London and her research is supported by the NIHR.

**Conflict of Interest** Disclosure The authors declare no conflict of interest.

### Competing Interest Statement

The authors have declared no competing interest.

### Funding Statement

The Developing Human Connectome Project was funded by the European Research Council under the European Union Seventh Framework Programme (FP/20072013)/ERC Grant Agreement no. 319456. This work was supported by the NIHR Biomedical Research Centres at Guys and St Thomas NHS Trust and the South London and Maudsley NHS Trust; the ESPRC/Wellcome Centre for Medical Engineering at Kings College London ((WT 203148/Z/16/Z); and the MRC Centre for Neurodevelopmental Disorders. The views expressed are those of the authors and not necessarily those of the NHS, the National Institute for Health Research or the Department of Health. The funders had no role in the design and conduct of the study; collection, management, analysis, and interpretation of the data; preparation, review, or approval of the manuscript; and decision to submit the manuscript for publication.
DB received support from a Wellcome Trust Seed Award in Science [217316/Z/19/Z]. LCG received support from the Comunidad de Madrid-Spain Support for R&D Projects [BGP18/00178]. H.C is an academic clinical lecturer in Clinical Genetics at Kings College London and her research is supported by the NIHR.

### Author Declarations

The study was approved by the United Kingdom Health Research Authority (Research Ethics Committee reference number: 14/LO/1169, IRAS 138070) and written informed consent was obtained from parents.

## References

1. Elliott LT, Sharp K, Alfaro-almagro F, Shi S, Miller KL, Douaud G, et al. Genome-wide association studies of brain imaging phenotypes in UK Biobank. 2018;

2. Hibar DP, Stein JL, Renteria ME, Arias-Vasquez A, Desrivières S, Jahanshad N, et al. Common genetic variants influence human subcortical brain structures. Nature [Internet]. 2015;520(7546):224–9. Available from: http://www.nature.com/doifinder/10.1038/nature14101

3. Satizabal CL, Adams HHH, Hibar DP, White CC, Knol MJ, Stein JL, et al. Genetic architecture of subcortical brain structures in 38,851 individuals. Nat Genet [Internet]. 2019;51(November). Available from: http://www.ncbi.nlm.nih.gov/pubmed/31636452

4. Xia K, Zhang J, Ahn M, Jha S, Crowley JJ, Szatkiewicz J, et al. Genome-wide association analysis identifies common variants influencing infant brain volumes. Transl Psychiatry. 2017;7(8):1–10.

5. Lamballais S, Jansen PR, Labrecque JA, Ikram MA, White T. Genetic scores for adult subcortical volumes associate with subcortical volumes during infancy and childhood. Hum Brain Mapp. 2021;42(6):1583–93.

6. Silbereis JC, Pochareddy S, Zhu Y, Li M, Sestan N. The Cellular and Molecular Landscapes of the Developing Human Central Nervous System. Neuron [Internet]. 2016;89(2):248. Available from: http://dx.doi.org/10.1016/j.neuron.2015.12.008

7. Hibar DP, Adams HHH, Jahanshad N, Chauhan G, Stein JL, Hofer E, et al. Novel genetic loci associated with hippocampal volume. Nat Commun. 2017;8.

8. Shepherd GMG. Corticostriatal connectivity and its role in disease. Nat Rev Neurosci. 2013;14(4):278–91.

9. Ellison-Wright I, Ellison-Wright Z, Bullmore E. Structural brain change in Attention Deficit Hyperactivity Disorder identified by meta-analysis. BMC Psychiatry. 2008;8:1–8.

10. Schmaal L, Veltman DJ, van Erp TGM, Sämann PG, Frodl T, Jahanshad N, et al. Subcortical brain alterations in major depressive disorder: findings from the ENIGMA Major Depressive Disorder working group. Mol Psychiatry [Internet]. 2016;21(6):806–12. Available from: http://www.nature.com/doifinder/10.1038/mp.2015.69

11. Van Erp TGM, Hibar DP, Rasmussen JM, Glahn DC, Pearlson GD, Andreassen OA, et al. Subcortical brain volume abnormalities in 2028 individuals with schizophrenia and 2540 healthy controls via the ENIGMA consortium. Mol Psychiatry. 2016;21(4):547–53.

12. Boardman JP, Counsell SJ, Rueckert D, Kapellou O, Bhatia KK, Aljabar P, et al. Abnormal deep grey matter development following preterm birth detected using deformation-based morphometry. Neuroimage. 2006;32(1):70–8.

13. Srinivasan L, Dutta R, Counsell SJ, Allsop JM, Boardman JP. Quantification of Deep Gray Matter in Preterm Infants at Term-Equivalent Age Using Manual Volumetry of 3-Tesla Magnetic Resonance Images. 2007;119(4).

14. Inder TE, Warfield SK, Wang H, Hu PS. Abnormal Cerebral Structure Is Present at Term in Premature Infants. 2005;115(2).

15. Boardman JP, Craven C, Valappil S, Counsell SJ, Dyet LE, Rueckert D, et al. A common neonatal image phenotype predicts adverse neurodevelopmental outcome in children born preterm. Neuroimage [Internet]. 2010;52(2):409–14. Available from: http://dx.doi.org/10.1016/j.neuroimage.2010.04.261

16. Edwards AD, Rueckert D, Smith SM, Abo Seada S, Alansary A, Almalbis J, et al. The Developing Human Connectome Project Neonatal Data Release. Front Neurosci. 022;16(May):1–14.

17. Hughes EJ, Winchman T, Padormo F, Teixeira R, Wurie J, Sharma M, et al. A dedicated neonatal brain imaging system. Magn Reson Med. 2017;78(2):794–804.

18. Makropoulos A, Robinson EC, Schuh A, Wright R, Fitzgibbon S, Bozek J, et al. The developing human connectome project: A minimal processing pipeline for neonatal cortical surface reconstruction. Neuroimage [Internet]. 2018;173(January):88–112. Available from: https://doi.org/10.1016/j.neuroimage.2018.01.054

19. Shi F, Yap PT, Wu G, Jia H, Gilmore JH, Lin W, et al. Infant brain atlases from neonates to 1- and 2-year-olds. PLoS One. 2011;6(4).

20. Andreas Schuh, Antonios Makropoulos, Emma C. Robinson, Lucilio Cordero-Grande, Emer Hughes, Jana Hutter, et al. Unbiased construction of a temporally consistent morphological atlas of neonatal brain development. bioRxiv. 2018;

21. Taoudi-Benchekroun Y, Christiaens D, Grigorescu I, Gale-Grant O, Schuh A, Pietsch M, et al. Predicting age and clinical risk from the neonatal connectome. Neuroimage [Internet]. 2022;257(May):119319. Available from: https://doi.org/10.1016/j.neuroimage.2022.119319

22. Patel H, Lee SH, Breen G, Menzel S, Ojewunmi O, Dobson RJB. The COPILOT Raw Illumina Genotyping QC Protocol. Curr Protoc. 2022;2(4):1–30.

23. McCarthy S, Das S, Kretzschmar W, Delaneau O, Wood AR, Teumer A, et al. A reference panel of 64,976 haplotypes for genotype imputation. Nat Genet. 2016;48(10):1279–83.

24. Auton A, Abecasis GR, Altshuler DM, Durbin RM, Bentley DR, Chakravarti A, et al. A global reference for human genetic variation. Nature. 2015;526(7571):68–74.

25. Choi SW, O’Reilly PF. PRSice-2: Polygenic Risk Score software for biobank-scale data. Gigascience. 2019;8(7):1–6.

26. Li J, Ji L. Adjusting multiple testing in multilocus analyses using the eigenvalues of a correlation matrix. Heredity (Edinb). 2005;95(3):221–7.

27. Lee PH, Anttila V, Won H, Feng YCA, Rosenthal J, Zhu Z, et al. Genomic Relationships, Novel Loci, and Pleiotropic Mechanisms across Eight Psychiatric Disorders. Cell. 2019;179(7):1469–1482.e11.

28. Medina-Gomez C, Felix JF, Estrada K, Peters MJ, Herrera L, Kruithof CJ, et al. Challenges in conducting genome-wide association studies in highly admixed multi-ethnic populations: the Generation R Study. Eur J Epidemiol. 2015;30(4):317–30.

29. Hibar DP, Stein JL, Renteria ME. Common genetic variants influence human subcortical brain structures. Nature [Internet]. 2015;520(7546):224–9. Available from: http://stacks.cdc.gov/view/cdc/31126/cdc_31126_DS1.pdf http://www.nature.com/nature/journal/vaop/ncurrent/full/nature14101.html?WT.ec_id=NATURE-20150122

30. Luo X, Guo X, Tan Y, Zhang Y, Garcia-Milian R, Wang Z, et al. KTN1 variants and risk for attention deficit hyperactivity disorder. Am J Med Genet Part B Neuropsychiatr Genet. 2020;(March):234–44.

31. Kang HJ, Kawasawa YI, Cheng F, Zhu Y, Xu X, Li M, et al. Spatio-temporal transcriptome of the human brain. Nature. 2011;478(7370):483–9.

32. Liu XA, Rizzo V, Puthanveettil S V. Pathologies of axonal transPort in neurodegenerative diseases. Transl Neurosci. 2012;3(4):355–72.

33. Xu B, Jia T, Macare C, Banaschewski T, Bokde ALW, Bromberg U, et al. Impact of a Common Genetic Variation Associated With Putamen Volume on Neural Mechanisms of Attention-Deficit/Hyperactivity Disorder. J Am Acad Child Adolesc Psychiatry. 2017;56(5):436–444.e4.

34. Choi SW, Mak TSH, O’Reilly PF. Tutorial: a guide to performing polygenic risk score analyses. Nat Protoc [Internet]. 2020;15(9):2759–72. Available from: http://dx.doi.org/10.1038/s41596-020-0353-1

35. Purcell SM, Wray NR, Stone JL, Visscher PM, O’Donovan MC, Sullivan PF, et al. Common polygenic variation contributes to risk of schizophrenia and bipolar disorder. Nature. 2009;460(7256):748–52.

36. Giedd JN. Stuctural Magnetic Resonance Imaging of the Adolescent Brain. Ann N Y Acad Sci [Internet]. 2004;1021:77–85. Available from: https://doi.org/10.1196/annals.1308.009

37. Dima D, Modabbernia A, Papachristou E, Doucet GE, Agartz I, Aghajani M, et al. Subcortical volumes across the lifespan: Data from 18,605 healthy individuals aged 3–90 years. Hum Brain Mapp. 2021;(June 2020):452–69.

38. Gilmore JH, Schmitt JE, Knickmeyer RC, Smith JK, Lin W, Styner M, et al. Genetic and environmental contributions to neonatal brain structure: A twin study. Hum Brain Mapp. 2010;31(8):1174–82.

39. Jansen AG, Mous SE, White T, Posthuma D, Polderman TJC. What Twin Studies Tell Us About the Heritability of Brain Development, Morphology, and Function: A Review. Neuropsychol Rev. 2015;25(1):27–46.

40. Reus LM, Shen X, Gibson J, Wigmore E, Ligthart L, Adams MJ, et al. Association of polygenic risk for major psychiatric illness with subcortical volumes and white matter integrity in UK Biobank. Nat Publ Gr [Internet]. 2017;(February):1–8. Available from: http://dx.doi.org/10.1038/srep42140

41. Franke B, Stein JL, Ripke S, Anttila V, Hibar DP, van Hulzen KJE, et al. Genetic influences on schizophrenia and subcortical brain volumes: large-scale proof of concept. Nat Neurosci [Internet]. 2016;19(3):420–31. Available from: https://samba.huji.ac.il/+CSCO+1h756767633A2F2F6A6A6A2E616E676865722E70627A++/neuro/journal/v19/n3/full/nn.4228.html

42. Alnæs D, Kaufmann T, Van Der Meer D, Córdova-Palomera A, Rokicki J, Moberget T, et al. Brain Heterogeneity in Schizophrenia and Its Association with Polygenic Risk. JAMA Psychiatry. 2019;76(7):739–48.

43. Grama S, Willcocks I, Hubert JJ, Pardiñas AF, Legge SE, Bracher-Smith M, et al. Polygenic risk for schizophrenia and subcortical brain anatomy in the UK Biobank cohort. Transl Psychiatry [Internet]. 2020;10(1). Available from: http://dx.doi.org/10.1038/s41398-020-00940-0

44. Acosta H, Kantojärvi K, Tuulari JJ, Lewis JD, Hashempour N, Scheinin NM, et al. Sex-specific association between infant caudate volumes and a polygenic risk score for major depressive disorder. J Neurosci Res. 2020;98(12):2529–40.

45. Alemany S, Jansen PR, Muetzel RL, Marques N, El Marroun H, Jaddoe VWV, et al. Common Polygenic Variations for Psychiatric Disorders and Cognition in Relation to Brain Morphology in the General Pediatric Population. J Am Acad Child Adolesc Psychiatry. 2019;58(6):600–7.

46. Cullen H, Krishnan ML, Selzam S, Ball G, Visconti A, Saxena A, et al. Polygenic risk for neuropsychiatric disease and vulnerability to abnormal deep grey matter development. 2019;(October 2018):1–8.

47. Cullen H, Selzam S, Dimitrakopoulou K, Plomin R, Edwards AD. Greater genetic risk for adult psychiatric diseases increases vulnerability to adverse outcome after preterm birth. Sci Rep [Internet]. 2021;11(1):1–8. Available from: https://doi.org/10.1038/s41598-021-90045-5

