## Supplementary Information for "Common Genetic Variation Important in Early Subcortical Brain Development"

**This file includes:**

**Supplementary Tables**: Supplementary Tables S1-S3

**Supplementary Figures:** Supplementary Figure S1

**Supplementary References**

**Supplementary Table S1:** SNP-volume pairs considered for exploration in our neonatal cohort

| **Subcortical Brain Volume** | **SNP** | **Main adult study *P* value** (1) (2) | **Allele 1/**  **Allele 2** | **Allele 1 Frequency** | **Replicated in Biobank** (3) | **Biobank study**  ***P* value** | **Explored in our neonatal cohort?** |
| --- | --- | --- | --- | --- | --- | --- | --- |
| Amygdala | rs11111293 | 4.16 × 10^−10^ | T/C | 0.78 | No | NA | No |
| **Brainstem** | **rs11111090** | **3.70 × 10^−27^** | **A/C** | **0.52** | **Yes** | **3.6x10^-12^** | **Yes** |
| **Caudate nucleus** | **rs3133370** | **5.59 × 10^−14^** | **T/C** | **0.67** | **Borderline** | **1.3x10^-8^** | **Yes** |
| **Globus pallidus** | **rs2923447** | **4.88 × 10^−16^** | **T/G** | **0.59** | **Yes** | **4.2x10^-17^** | **Yes** |
| **Putamen** | **rs945270** | **5.02 × 10^−51^** | **C/G** | **0.58** | **Yes** | **4.0x10^-12^** | **Yes** |
| Thalamus | rs12600720 | 4.06 × 10^−10^ | C/G | 0.69 | No | NA | No |
| **Hippocampus** | **rs77956314** | **2.06 × 10^−25^** | **T/C** | **0.916** | **Yes** | **1.2x10^-11^** | **Yes** |
| **Hippocampus** | **rs61921502** | **1.94 × 10^−19^** | **T/G** | **0.846** | **Borderline** | **1.8x10^-9^** | **Yes** |

**Supplementary Table 1**. Information on the eight SNPs most robustly associated with subcortical brain volumes from studies (1) and (2). The table details the *P* value of association in the original adult study, allele 1 and 2 for the SNP in question, the allele frequency of allele 1 and details of the whether the SNP-volume association was replicated in (3). The nucleus accumbens is not included because this volume was not extracted using our neonatal atlases.

**Supplementary Table S2:** Publicly available Psychiatric Genomics Consortium GWAS summary statistics used for the computation of the GPSs in this study

| **Psychiatric Disorder** | **Reference** | **Year Published** | **Cases** | **Controls** | **Sample Size** |
| --- | --- | --- | --- | --- | --- |
| ADHD | Demontis et al. (4) | 2019 | 20183 | 35191 | 55374 |
| ASD | Grove et al. (5) | 2019 | 18381 | 27969 | 46350 |
| Bipolar Disorder | Mullins et al. (6) | 2021 | 41917 | 371549 | 413466 |
| Schizophrenia | Trubetskoy et al. (7) | 2022 | 53386 | 77258 | 130644 |
| Major Depressive Disorder | Howard et al. (8) | 2019 | 246363 | 561190 | 807,553 |
| Cross Disorder | Lee et al. (9) | 2019 | 232964* | 494162* | 727126* |

### **Supplementary Table S2**. Publicly available Psychiatric Genomics Consortium GWAS summary statistics used for the computation of the GPSs in this study. ADHD – Attention Deficit Hyperactivity Disorder, ASD – Autism Spectrum Disorder. * Actual cohort smaller as data from 23andMe were excluded.

**Supplementary Table S3a**: SNP-volume results for the European-Asian ancestry (n=258) cohort

| Volume | Marker | Allele 1 | Allele 2 | Allele 2 AF | Allele 1 AF | Rsq | European-Asian ancestry cohort (n=258) | | |
| --- | --- | --- | --- | --- | --- | --- | --- | --- | --- |
|  |  |  |  |  |  |  | *P* value | β | SE |
| **Putamen** | **rs945270** | **C** | **G** | **0.51713** | **0.48287** | **0.97845** | **1.89x10^-3^** | **0.1251** | **0.0399** |
| Pallidum | rs2923447 | **G** | T | 0.58867 | 0.41133 | 0.98606 | 0.4956 | 0.0295 | 0.0432 |
| Caudate | rs3133370 | **C** | T | 0.62460 | 0.3754 | Genotyped | 0.278 | -0.0489 | 0.0450 |
| *Brainstem* | *rs11111090* | *C* | *A* | *0.62688* | *0.37312* | *0.9914* | *0.0293* | *-0.0708* | *0.0323* |
| Hippocampus | rs77956314 | **C** | T | 0.92469 | 0.07531 | 0.98433 | 0.168 | 0.0600 | 0.0435 |
| *Hippocampus* | *rs61921502* | *G* | *T* | *0.88988* | *0.11012* | *Genotyped* | *0.0202* | *-0.1012* | *0.0433* |

**Supplementary Table S3a.** Results of association analyses for the six selected SNP-volume pairs in our cohort of 258 European-Asian ancestry neonates. Allele 1 is the coded allele; Allele 2 is the non-coded allele. Standardized beta coefficients (β) and standard errors (SE) are given with respect to Allele 1. The table provides raw *P* values. Results surviving Bonferroni-correction (*P* <0.0083) are indicated in bold. Results indicating nominal significance (*P* <0.05) are indicated in italics.

**Supplementary Table S3b:** SNP-volume results for the full mixed-ancestry (n=418) cohort

| Volume | Marker | Allele 1 | Allele 2 | Allele 2 AF | Allele 1 AF | Rsq | Full mixed-ancestry cohort (n=418) | | |
| --- | --- | --- | --- | --- | --- | --- | --- | --- | --- |
|  |  |  |  |  |  |  | *P* value | β | SE |
| **Putamen** | **rs945270** | **C** | **G** | **0.51713** | **0.48287** | **0.97845** | **4.16x10^-4^** | **0.1223** | **0.0343** |
| Pallidum | rs2923447 | **G** | T | 0.58867 | 0.41133 | 0.98606 | 0.4490 | 0.0250 | 0.0329 |
| Caudate | rs3133370 | **C** | T | 0.62460 | 0.3754 | Genotyped | 0.5941 | -0.0186 | 0.0348 |
| Brainstem | rs11111090 | **C** | A | 0.62688 | 0.37312 | 0.9914 | 0.1297 | -0.0429 | 0.0283 |
| Hippocampus | rs77956314 | **C** | T | 0.92469 | 0.07531 | 0.98433 | 0.2298 | 0.0405 | 0.0337 |
| *Hippocampus* | *rs61921502* | *G* | *T* | *0.88988* | *0.11012* | *Genotyped* | *0.0375* | *-0.0705* | *0.0338* |

**Supplementary Table S3b.** Results of association analyses for the six selected SNP-volume pairs in our cohort of 418 mixed-ancestry neonates. Allele 1 is the coded allele; Allele 2 is the non-coded allele. Standardized beta coefficients (β) and standard errors (SE) are given with respect to Allele 1. The table provides raw *P* values. Results surviving Bonferroni-correction (*P* <0.0083) are indicated in bold. Results indicating nominal significance (*P* <0.05) are indicated in italics.

**Supplementary Figure 1:** Heatmaps for association between adult subcortical genome-wide polygenic scores and neonatal brain volume

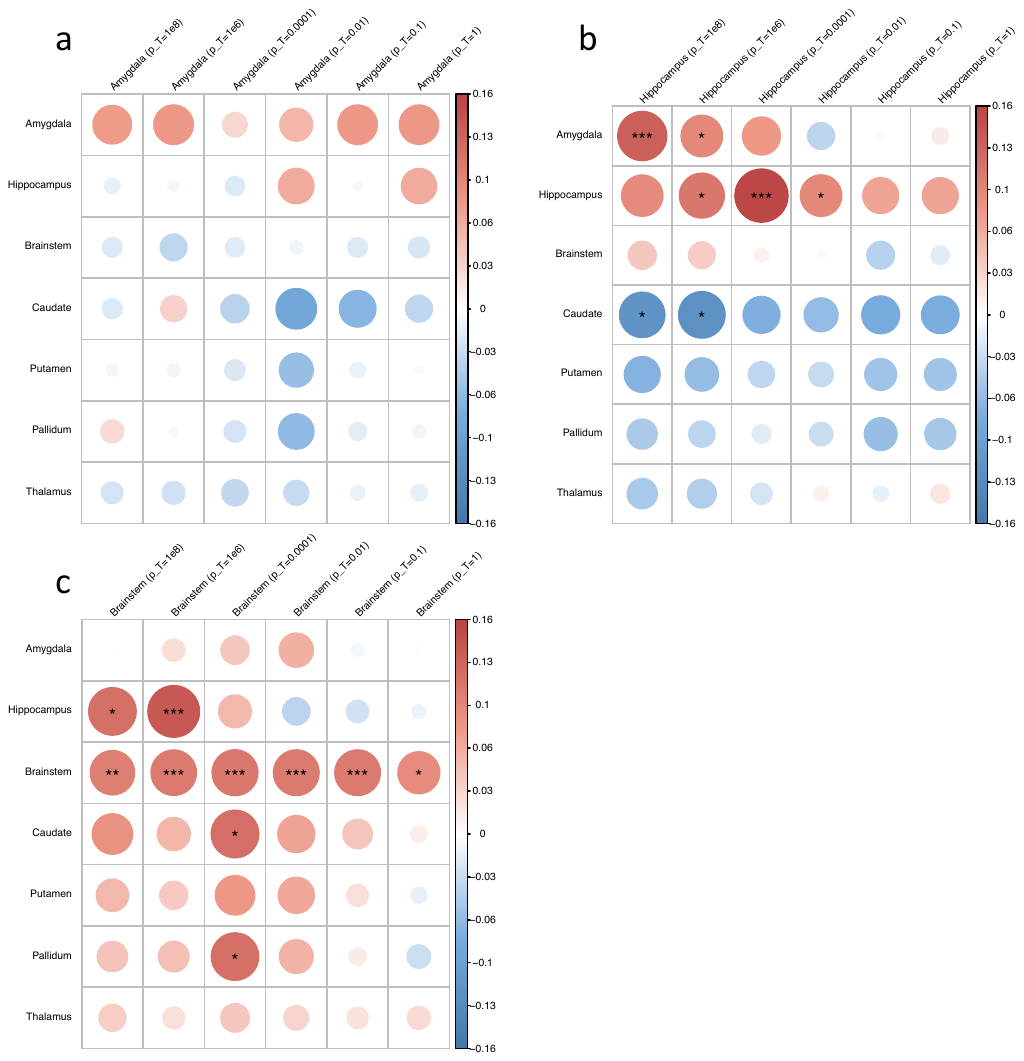

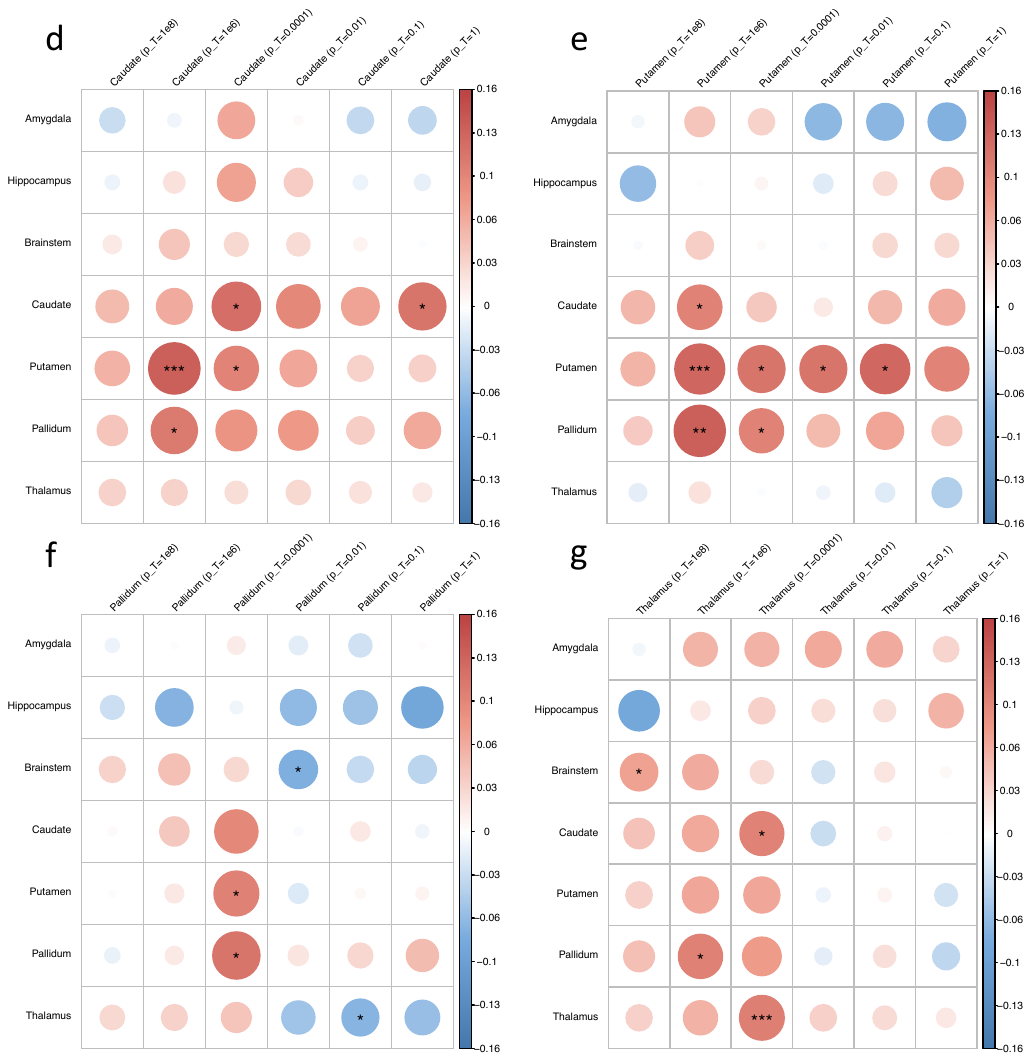

**Supplementary Figure 1.** Heatmaps for the associations between GPSs for seven adult subcortical brain volumes and the corresponding neonatal brain volumes. Each heatmap corresponds the GPSs for a different adult subcortical brain volume: Amygdala (a), Hippocampus (b), Brainstem (c), Caudate (d), Putamen (e), Pallidum (f) and Thalamus (g). The horizontal axis gives the GPS at the six GWAS *P*-value thresholds (p_T_ = 1x10^-8^, 1x10^-6^, 0.0001, 0.01, 0.1 and 1) and the vertical axis the seven neonatal subcortical brain volumes. **P* value < 0.05; ** *P* value <0.01; ***P value < 4.17x10^-3^.

**Supplementary References**

1.	Satizabal CL, Adams HHH, Hibar DP, White CC, Knol MJ, Stein JL, et al. Genetic architecture of subcortical brain structures in 38,851 individuals. Nat Genet [Internet]. 2019;51(November). Available from: http://www.ncbi.nlm.nih.gov/pubmed/31636452

2.	Hibar DP, Adams HHH, Jahanshad N, Chauhan G, Stein JL, Hofer E, et al. Novel genetic loci associated with hippocampal volume. Nat Commun. 2017;8.

3.	Elliott LT, Sharp K, Alfaro-almagro F, Shi S, Miller KL, Douaud G, et al. Genome-wide association studies of brain imaging phenotypes in UK Biobank. 2018;

4.	Demontis D, Walters RK, Martin J, Mattheisen M, Als TD, Agerbo E, et al. Discovery of the first genome-wide significant risk loci for attention deficit/hyperactivity disorder. Nat Genet. 2019;51(1):63–75.

5.	Grove J, Ripke S, Als TD, Mattheisen M, Walters RK, Won H, et al. Identification of common genetic risk variants for autism spectrum disorder. Nat Genet. 2019;51(3):431–44.

6.	Mullins N, Forstner AJ, O’Connell KS, Coombes B, Coleman JRI, Qiao Z, et al. Genome-wide association study of more than 40,000 bipolar disorder cases provides new insights into the underlying biology. Nat Genet. 2021;53(6):817–29.

7.	Trubetskoy V, Pardiñas AF, Qi T, Panagiotaropoulou G, Awasthi S, Bigdeli TB, et al. Mapping genomic loci implicates genes and synaptic biology in schizophrenia. Nature. 2022;604(7906):502–8.

8.	Howard DM, Adams MJ, Clarke TK, Hafferty JD, Gibson J, Shirali M, et al. Genome-wide meta-analysis of depression identifies 102 independent variants and highlights the importance of the prefrontal brain regions. Nat Neurosci [Internet]. 2019;22(3):343–52. Available from: http://dx.doi.org/10.1038/s41593-018-0326-7

9.	Lee PH, Anttila V, Won H, Feng YCA, Rosenthal J, Zhu Z, et al. Genomic Relationships, Novel Loci, and Pleiotropic Mechanisms across Eight Psychiatric Disorders. Cell. 2019;179(7):1469-1482.e11.
